# Plasma Phosphorylated-tau 217 Reshapes Diagnostic Classification in a Real-World Memory Clinic Cohort

**DOI:** 10.64898/2026.06.20.26356107

**Authors:** Nora Maerean, Sofia Litchev, George R. Jackson, Joseph S. Kass, Valory N. Pavlik, Chi-Ying R. Lin

**Affiliations:** Alzheimer’s Disease and Memory Disorders Center, Department of Neurology, Baylor College of Medicine, Houston, Texas, USA; Parkinson’s Disease Center and Movement Disorders Clinic, Department of Neurology, Baylor College of Medicine, Houston, Texas, USA

## Abstract

**Background:** Plasma phosphorylated tau-217 (ptau217) has demonstrated accuracy exceeding 90% for Alzheimer’s disease (AD) diagnosis. While its diagnostic validity has been established, its real-world clinical utility in altering or corroborating clinician diagnoses remains less understood. This study evaluates the impact of plasma ptau217 on diagnostic reclassification in a memory disorders clinic.

**Methods:** We conducted a retrospective chart review of 100 patients evaluated for memory impairment who subsequently underwent plasma biomarker testing. Initial clinical diagnoses were established using the National Institute of Neurological and Communicative Disorders and Stroke/Alzheimer Disease and Related Disorders Association criteria, neuropsychological testing, and brain magnetic resonance imaging without knowledge of plasma biomarker results. Follow-up diagnoses were assigned after the availability of plasma ptau217 or Precivity AD2 results. Diagnostic reclassification was evaluated under two p-tau217 classification schemes to determine the AD etiology: a binary cutoff with lower threshold (AD = ptau217 > 0.18) and a three-level cutoff incorporating a higher threshold (AD = ptau217 > 0.325). Reclassification matrices and McNemar’s tests were used to assess changes in diagnosis. Subgroup analyses were conducted according to apolipoprotein E (*APOE) ε4* carrier status.

**Results:** The overall diagnosis reclassification rates were 37.8% (χ^2^ (1) = 10.81, *p = 0.001)* and 46.9% (χ^2^ (1) = 29.76; *p < 0.001)* using lower and higher threshold analyses, respectively. With the higher threshold (ptau217 > 0.325), reclassification among *APOE ε4* carriers occurred in both directions at similar frequencies, without evidence of a net shift toward AD or non-AD (42% from AD to non-AD, 43% from non-AD to AD, χ² (1) = 2.77 *p = 0.096*). Among *ε4* non-carriers, reclassification was significantly asymmetric toward non-AD (66% from AD to non-AD, 14% from non-AD to AD, χ² (1) = 16.41, *p < 0.001*), suggesting ptau217 may be identifying diagnostically heterogeneous cases in which the clinical presentation resembles AD but instead reflects an alternative or co-morbid etiologies.

**Conclusion:** Plasma ptau217 meaningfully influences diagnostic decisions in a memory clinic setting and may be particularly valuable among *APOE ε4* non-carriers, where biomarker-informed evaluation frequently shifted diagnoses away from AD.

**TAKE-HOME POINTS:**

- Phosphorylated tau-217 meaningfully influences clinical diagnosis in a real-world memory disorders clinic.
- Higher phosphorylated tau-217 threshold favors diagnostic reclassification from Alzheimer’s to non-Alzheimer’s disease etiologies.
- The clinical impact of phosphorylated tau-217 on the etiological diagnosis change is greatest among *APOE ε4* non-carriers.

## Introduction

Alzheimer’s disease (AD) has become increasingly prevalent as the world’s aging population grows. In 2024, approximately 6.9 million Americans over the age of 65 had AD (Alzheimer’s Association, 2024). In the past, the diagnosis of AD had been based on clinical characteristics and neuropsychological testing only;^1^ however, with the development of neuroimaging and fluid biomarkers, diagnostic criteria have shifted towards pathologic changes.^2^ In 2024, the Alzheimer’s Association Workgroup 2024 defined AD as an abnormality in one of the defined Core 1 biomarkers.^2^ These include abnormalities in amyloid positron emission tomography (PET) scans, cerebrospinal fluid (CSF) biomarkers, or plasma assays with accuracy equivalent to already approved CSF assays.^2^ These Core 1 biomarkers can identify the presence of AD pathology in both symptomatic and asymptomatic individuals.^2^

A few challenges currently exist in the diagnosis of AD. Firstly, symptoms of dementias overlap and thus can lead to misclassification or misdiagnosis.^3–5^ The increased use of blood-based biomarkers may also result in more asymptomatic individuals being labeled as having Alzheimer’s disease pathology despite uncertainty regarding whether they will follow the typical clinical trajectory toward mild cognitive impairment or dementia later in life.^6^

Although traditional immunohistochemistry approaches to studying tau phosphorylation using conformation-specific antibodies have been used for decades, in recent years, specific tau phosphorylation sites were identified to correlate with severity of neuronal cytopathology in AD^7^. Phosphorylated tau-217 (ptau217) has emerged as a promising blood-based biomarker for AD, with numerous studies demonstrating strong diagnostic performance and concordance with other established AD.^8,9^ Mass spectrometry based approaches have identified ptau217 as highly prevalent in AD brain banks and less enriched on other tauopathies.^10^ In addition, the Precivity AD2 blood tests, which contains %ptau217 and amyloid beta 42/40, have also demonstrated strong clinical validity for AD diagnosis.^11^ Furthermore, pTau-217 has been shown to have a low misclassification rate.^8^ These accuracies are similar to other CSF biomarkers in finding amyloid and tau abnormalities.^12^ However, while many studies have established that ptau217 is a highly accurate biomarker that can be used to diagnose AD, fewer studies have been conducted to demonstrate its utility in the clinical settings. The purpose of this study is to evaluate the real-world application of blood-based ptau217 as a diagnostic biomarker of AD. Specifically, we aimed to investigate whether the use of ptau217 can meaningfully alter or corroborate the initial clinic diagnosis in a memory clinic of an academic medical center.

## Methods

### Study design and participants

We conducted a retrospective chart review study with patients identified through the electronic medical record at the Baylor College of Medicine Alzheimer’s Disease and Memory Disorders Center (ADMDC) from June 1, 2025, to December 31, 2025. The study was approved by the Baylor College of Medicine Institutional Review Board. Participants were identified through the electronic medical record and screened from individuals aged 50 years or older who underwent evaluation for memory impairment and had available plasma biomarker testing (ptau217 or PrecivityAD2) as part of their clinical diagnostic workup. To reflect the real-world clinical population encountered in a tertiary memory disorders clinic, patients with both neurodegenerative and non-neurodegenerative causes of cognitive impairment were eligible for inclusion. One hundred eligible patients were included in the final cohort. Initial cognitive diagnoses were established by ADMDC neurologists based on history, clinical symptoms, comprehensive neuropsychological testing, and brain MRI without ptau217, CSF studies, or amyloid PET; AD diagnosis was assigned according to the National Institute of Neurological and Communicative Disorders and Stroke/Alzheimer Disease and Related Disorders Association (NINCDS/ADRDA) criteria.^13^ Non-AD diagnoses include frontotemporal dementia, vascular dementia, Lewy body dementia, limbic-predominant age-related TDP-43 encephalopathy, primary age-related tauopathy, cognitive changes driven by other factors, including multiple sclerosis, neurofibromatosis type 1, traumatic brain injury, paraneoplastic syndrome, and mood as well as psychiatric disorders, including depression and attention deficit and hyperactive disorder. A follow-up diagnosis was assigned after the availability of plasma ptau217 or/and Precivity AD2 results. Diagnostic reclassification was assessed by comparing the follow-up biomarker-driven diagnosis with the initial clinical diagnosis. All cases were reviewed in the ADMDC multidisciplinary diagnostic consensus conference, where comprehensive clinical, neuropsychological, neuroimaging, and biomarker data were discussed to corroborate diagnostic classifications.

### Demographics and measures

Our data collection included the following demographic data: age at time of data collection, age at symptom onset, and apolipoprotein E (*APOE*) testing results. Mental status exam items collected include Mini-Mental State Examination (MMSE) scores (two scores per participant, by both serial 7s and backward spelling), and clock drawing test score. Blood-based biomarker testing consisted of plasma phosphorylated tau-217 (ptau217) measured using the Labcorp Lumipulse assay and the PrecivityAD2 test (C2N Diagnostics). We applied two approaches to determine the blood-based biomarker-informed diagnosis of AD. First, a binary classification approach defined AD as ptau217 > 0.18, consistent with the Labcorp Lumipulse assay interpretation,^14^ or a positive “Amyloid Probability Score 2” on the PrecivityAD2 test.^11^ Second, a three-level classification approach incorporated a higher ptau217 threshold, with ptau217 > 0.325 considered supportive of AD pathology based on the Labcorp Lumipulse assay reference ranges^15^. For PrecivityAD2, classification was based on the manufacturer-reported “Amyloid Probability Score 2” interpretation categories.

### Statistical analysis

The impact of ptau-217 results on clinical diagnosis was evaluated using a reclassification matrix. McNemar’s test was used to evaluate asymmetry in paired diagnostic classifications before and after incorporation of ptau217 results. The test specifically compares discordant pairs (AD → non-AD vs non-AD → AD, examples in **Supplemental Table 1**). For the three-level ptau217 categorization, intermediate values were collapsed into the non-AD category to permit binary comparison with the initial clinical diagnosis. Thus, McNemar’s test was then also applied to evaluate asymmetry in paired diagnostic classifications before and after incorporation of ptau217 results. Additionally, we performed exploratory subgroup analyses stratified by *APOE ε4* carrier status. Within each stratum, McNemar’s test was used to assess asymmetry in paired diagnostic reclassification after incorporation of ptau217 results. Differences in reclassification patterns by *APOE* status were evaluated descriptively.

### Standard Protocol Approvals, Registrations, and Patient Consents

Patients or their legally authorized representatives consented to participate in the study.

### Data Availability

De-identified data not published in this article will be made available by request from investigators.

## Results

### Demographics

We evaluated a total of 100 patients in the ADMDC clinic, of which 98 had either ptau217 (n = 75) or Precivity AD2 testing done (n = 23) and were used for analysis. Our patient population had a mean age of 73.06 ± 8.85 year, with mean disease duration at time of data collection 4.76 ± 3.72. Participants had an average of 16.07 ± 3.00 years of education. Mean MMSE 7’s score was 23.91 ± 4.86 and mean MMSE spelling score was 25.51 ± 3.98. *APOE* genotype testing showed that 31 (43.66%) patients were *APOE ε4* carriers (n= 71 patients who received *APOE* testing). Of these individuals, 25 (35.71% of total) were heterozygous while 6 (8.45%) were homozygous carriers. Forty patients (56.34%) were *APOE ε4* non-carriers. Full demographics and clinical characteristics are listed in **Table 1**.

**Table 1:** Demographic and clinical characteristics of the study cohort.

|  |  |
| --- | --- |
| Age (mean ± SD) | 73.43 ± 8.74 |
| Age onset (mean ± SD) | 68.57 ± 10.43 |
| Disease duration (mean ± SD) | 4.76 ± 3.72 |
| Education (mean ± SD) | 16.07 ± 3.00 |
| Sex (F:M) | 36:34 |
| MMSE 7's (mean ± SD) | 23.91 ± 4.86 |
| MMSE spelling (mean ± SD) | 25.51 ± 3.98 |
| *APOE* ε4 non-carriers* | 40 (56.33%) |
| Heterozygous *APOE* ε4 carriers** | 25 (35.21%) |
| Homozygous *APOE* ε4 carriers | 6 (8.45%)* |

\* *APOE* ε4 non-carriers includes $\epsilon$ 2/3 (n = 4) and $\epsilon$ 3/3 (n = 36) genotypes
\*\* Heterozygous *APOE* ε4 carriers are $\epsilon$ 3/4 (n = 22) and $\epsilon$ 2/4 (n = 3) genotypes
\*\*\* These numbers reflect the distribution in *APOE* genotype among patients who had *APOE* testing done (n = 71).

### Overall distribution of initial clinical impression to final diagnoeis

Before applying binary or three-level ptau217 cutoff analyses, we first examined the overall concordance between clinicians’ initial impressions and biomarker-informed final diagnoses across the full cohort (**Table 2**). Among 98 patients evaluated for cognitive and behavioral disorders, 61 (62.2%) had a final diagnosis that matched the initial clinical impression. Specifically, 47 patients (48.0%) retained an AD diagnosis, while 14 patients (14.3%) retained a non-AD diagnosis. Diagnostic discordance occurred in 37 patients (37.8%). Of these, 29 patients (29.6%) were initially considered to have AD but were subsequently reclassified to non-AD etiologies after biomarker integration, whereas 8 patients (8.2%) were initially considered non-AD but were later reclassified as AD. These findings suggest that incorporation of plasma ptau217 and/or PrecivityAD2 testing more frequently shifted diagnostic impressions away from AD than toward AD in this memory clinic population. The non-AD etiologies identified among reclassified participants were heterogeneous (**Table 2**), underscoring the complexity of differential diagnosis in real-world cognitive neurology practice.

**Table 2:** Initial clinical diagnosis versus final diagnosis established by incorporating the etiological information from blood-based biomarkers.

| Initial impression | Final diagnosis | N (%) |
| --- | --- | --- |
| AD | AD | 47 (48.0%) |
| AD | Non-AD diagnosis<sup>1</sup> | 29 (29.6%) |
| Non- AD diagnosis<sup>2</sup> | AD | 8 (8.2%) |
| Non-AD diagnosis<sup>3</sup> | Non-AD diagnosis<sup>4</sup> | 14 (14.3%) |

AD: Alzheimer's disease. Overall concordance between initial clinical impression and final diagnosis was 62.2%.
<sup>1</sup> Patients' final diagnoses include frontotemporal dementia, vascular dementia, primary age-related tauopathy, cognitive changes driven by psychiatric disorder or multiple sclerosis, neurofibromatosis type 1, or not meeting cognitive impairment criteria on neuropsychological testing
<sup>2</sup> Patients' initial diagnostic impression include VaD, DLB, ADHD symptomatology, FTD, anti-P/Q voltage dependent channel antibodies
<sup>3</sup> Patients' initial diagnostic impression include Mood disorder and traumatic brain injury
<sup>4</sup> Patients' final diagnoses include Mood disorder and traumatic brain injury

### Lower threshold analysis (ptau217 > 0.18 as AD)

We developed a reclassification matrix representing initial impression and final diagnosis (**Table 3**) using a single cutoff value for ptau-217 results. In this classification scheme, patients were designated as having AD if they had a plasma ptau217 level > 0.18 or a positive PrecivityAD2 result, as determined by the manufacturer-defined “Amyloid Probability Score 2” classification. **Table 3** represents the reclassification matrix using these criteria. We found an overall diagnostic reclassification in 37.8% of patients evaluated for memory loss (n = 98). Reclassification occurred both in the direction towards AD and towards non-AD diagnosis. Among patients whose initial impression was Alzheimer’s Disease (n = 76), 38.2% were reclassified to non-AD etiology (n = 29). Among patients who had an initial impression of non-AD etiology (n = 22), 36.4% were given a final etiological diagnosis of AD (n = 8). We performed a McNemar test to assess whether the use of ptau-217 after first clinical impression significantly influences diagnostic change, which demonstrated significant asymmetry in diagnostic reclassification after incorporation of ptau217 results (χ^2^ (1) = 10.81, *p = 0.001*), indicating that diagnostic changes were more likely to occur in the direction of AD → non-AD than non-AD → AD.

**Table 3:** Diagnostic reclassification under binary and three-level plasma biomarker classification schemes.

| **Initial diagnosis** | **Diagnosis after single cutoff<sup>1</sup>** |  |
| --- | --- | --- |
|  | **AD** | **Non-AD diagnosis<sup>2</sup>** |
| **AD** | 47 | 29 |
| **Non-AD diagnosis** | 8 | 14 |

|  | **Diagnosis after three-level categorization<sup>3</sup>** |  |
| --- | --- | --- |
|  | **AD** | **Non-AD diagnosis<sup>4</sup>** |
| **AD** | 34 | 42 |
| **Non-AD diagnosis** | 4 | 18 |

<sup>1</sup> AD etiology is defined as pTau217 > 0.18 or a positive PrecivityAD2 test result<sup>2</sup> Non-AD etiology is designated for patients who have pTau217 < 0.18 or a negative PrecivityAD2 test result<sup>3</sup> AD etiology is defined as pTau217 > 0.325 or a positive PrecivityAD2 test result<sup>4</sup> Non-AD etiology is designated for patients who have pTau217 < 0.325 or a negative PrecivityAD2 test result

### Higher threshold analysis (ptau217 > 0.325 as AD)

Our second reclassification table used a three-level classification that uses a higher cutoff for detection of AD (ptau217 > 0.325). Patients who met the criteria ptau217 > 0.325 or positive PrecivityAD2 received a final etiological diagnosis of AD. Below is the reclassification matrix table (**Table 3**). The overall reclassification rate was 46.9%. 55.3% of individuals with an initial impression of AD etiology (n = 76) were reclassified to non-AD etiology (n = 42), while 18.2% of individuals within the non-AD category (n = 22) had a change in diagnosis to AD etiology (n = 18). McNemar’s test demonstrated a significant asymmetry in diagnostic reclassification after incorporation of the higher ptau217 threshold (χ^2^ (1) = 29.76; *p < 0.001*), indicating that diagnostic changes occurred more frequently in the direction of AD → non-AD than non-AD → AD when the more stringent cutoff was applied.

### *APOE* carrier status effect

To further evaluate whether *APOE ε4* carrier status influenced the directionality of biomarker-informed diagnostic reclassification, we performed subgroup analyses among participants who underwent both *APOE* genotyping and biomarker testing (n = 70). Within *APOE ε4* carriers (n = 31), incorporation of plasma ptau217 and/or Precivity AD2 results did not produce significant directional asymmetry in diagnostic change. Using the single cutoff with binary classification (ptau217 > 0.18 = AD, < 0.18 = non-AD), reclassification occurred in both directions, with 6 individuals initially classified as non-AD subsequently reclassified to AD, while 4 individuals initially classified as AD were reclassified to non-AD. McNemar’s testing within this subgroup was not statistically significant (χ^2^ (1) = 0.10, *p = 0.75*), suggesting that in *APOE ε4* carriers, plasma p-tau217 did not systematically shift the diagnostic classification toward or away from AD. In contrast, among *APOE ε4* non-carriers (n = 39), biomarker-informed diagnostic change was markedly asymmetric. Only 1 individual was reclassified from non-AD to AD (14%), whereas 18 individuals were reclassified from AD to non-AD following biomarker incorporation (56%). This directional shift was statistically significant (χ² (1) = 13.47, *p < 0.001*), indicating that among *ε4* non-carriers, plasma ptau217 more frequently redirected diagnostic impressions away from AD and toward alternative etiologies. Reclassification matrices for both *APOE ε4* carriers and non-carriers are shown in **Table 4**. Our results using two-point cutoff with ptau217 threshold set up as 0.325 yielded the similar findings: Among APOE ε4 carriers, reclassification was symmetric with 42% from AD to non-AD and 43% from non-AD to AD (χ²(1) = 2.77, *p = 0.096*, **Table 5**). Among ε4 non-carriers, reclassification was significantly asymmetric toward non-AD, with 66% from AD to non-AD and 14% from non-AD to AD (χ² (1) = 16.41, *p < 0.001,* **Table 5**).

**Table 4:** Diagnostic reclassification following plasma biomarker testing according to *APOE ε4* carrier status with AD etiology defined as ptau217 > 0.18.

| $\epsilon 4$ carriers | | |
| --- | --- | --- |
| Initial diagnosis | Follow-up diagnosis |  |
|  | AD<sup>1</sup> | Non-AD diagnosis<sup>2</sup> |
| AD | 20 | 4 |
| Non-AD diagnosis | 6 | 1 |

| Non-$\epsilon 4$ carriers | | |
| --- | --- | --- |
| Initial diagnosis | Follow-up diagnosis |  |
|  | AD | Non-AD diagnosis |
| AD | 14 | 18 |
| Non-AD diagnosis | 1 | 6 |

<sup>1</sup> AD etiology is defined as pTau217 > 0.18 or a positive PrecivityAD2 test result
<sup>2</sup> Non-AD etiology is designated for patients who have pTau217 < 0.18 or a negative PrecivityAD2 test result

**Table 5:** Diagnostic reclassification following plasma biomarker testing according to *APOE ε4* carrier status with AD etiology defined as ptau217 > 0.325.

| $\epsilon 4$ carriers | | |
| --- | --- | --- |
| Initial diagnosis | Follow-up diagnosis |  |
|  | AD<sup>1</sup> | Non-AD diagnosis<sup>2</sup> |
| AD | 14 | 10 |
| Non-AD diagnosis | 3 | 4 |

| Non-$\epsilon 4$ carriers | | |
| --- | --- | --- |
| Initial diagnosis | Follow-up diagnosis |  |
|  | AD | Non-AD diagnosis |
| AD | 11 | 21 |
| Non-AD diagnosis | 1 | 6 |

<sup>1</sup> AD etiology is defined as pTau217 > 0.325 or a positive PrecivityAD2 test result <sup>2</sup> Non-AD etiology is designated for patients who have pTau217 < 0.325 or a negative PrecivityAD2 test result

## Discussion

Plasma p-tau217 has emerged as one of the most diagnostically accurate blood-based biomarkers for Alzheimer’s disease (AD), with multiple studies demonstrating performance approaching established CSF and amyloid PET biomarkers.^8,9,12^ However, diagnostic accuracy alone does not fully capture clinical utility. In real-world practice, clinicians must determine how biomarker data meaningfully alter diagnostic reasoning beyond initial history, neuropsychological testing, and neuroimaging. Our study addresses this translational gap by demonstrating that plasma p-tau217 and/or Precivity AD2 testing substantially influenced diagnostic reclassification in a specialized memory disorders clinic, with approximately 38% to 47% of patients experiencing a change in etiological classification depending on the biomarker threshold applied. These findings suggest that blood-based biomarkers are not merely confirmatory tools but may meaningfully reshape diagnostic impressions in clinically complex populations.

A key finding of this study is that the direction and magnitude of diagnostic reclassification were strongly influenced by ptau217 threshold selection. Under a binary cutoff (> 0.18), reclassification occurred in both directions, suggesting that plasma p-tau217 can both support an initial AD diagnosis and identify patients whose presentations may warrant reconsideration toward non-AD etiologies. In contrast, the more stringent two-point cutoff (> 0.325) produced substantially greater directional asymmetry toward AD to non-AD reclassification. This pattern suggests that higher ptau217 thresholds may function more conservatively, potentially increasing specificity for biologically defined AD while reducing false-positive clinical impressions of AD in diagnostically heterogeneous cases. Of note, this should not be interpreted as reduced utility of ptau217 at higher thresholds, but rather as a shift in clinical function from broader detection toward greater rule-out value. At the same time, some patients with clinically suspected AD but lower ptau217 values may still represent biologically early or evolving AD in whom plasma biomarkers have not yet fully converted, highlighting an important temporal limitation of cross-sectional, single-timepoint biomarker interpretation.^16^

The *APOE ε4* subgroup findings further refine the clinical relevance of plasma p-tau217. Among *APOE ε4* carriers, biomarker incorporation produced relatively symmetric and statistically nonsignificant reclassification, with similar proportions of patients reclassified from AD to non-AD and from non-AD to AD. These findings indicate bidirectional diagnostic refinement rather than confirmation of the initial clinical impression or a systematic shift toward either diagnostic category.. By contrast, among *ε4* non-carriers, ptau217 more frequently shifted diagnoses away from AD, often toward alternative etiologies. This finding suggests that plasma p-tau217 may be particularly valuable in diagnostically ambiguous populations where baseline clinical suspicion of AD is less genetically reinforced. In this context, ptau217 may function less as a simple AD confirmation tool and more as a diagnostic refinement instrument that helps identify patients whose cognitive syndromes phenotypically resemble AD but may instead reflect vascular, frontotemporal, psychiatric, or mixed-pathology processes.^17–19^ In this setting, plasma p-tau217 may serve less as a confirmatory AD biomarker and more as a diagnostic refinement tool that improves etiologic specificity and reduces potential overdiagnosis of AD. This distinction may become increasingly important as blood-based biomarkers are incorporated into treatment pathways for anti-amyloid therapies and clinical trial participation, where accurate identification of underlying AD pathology has direct implications for patient selection, prognostication, and counseling.^20^ Furthermore, these findings support emerging precision medicine frameworks in which the clinical utility of biomarkers is interpreted in the context of an individual’s genetic background and baseline disease likelihood rather than as a uniform diagnostic test applied equally across all patients.^21^ From a clinical perspective, these data suggest that the greatest diagnostic yield from plasma p-tau217 may occur in patients whose clinical presentation is suggestive but not unequivocally consistent with AD, particularly among *APOE* ε4 non-carriers. Our findings reinforce that plasma p-tau217 should be integrated within comprehensive clinical contexts rather than used in isolation. Biomarker positivity or negativity alone may not fully capture co-pathology, disease stage, or longitudinal trajectory.

Our study has several limitations. First, this was a retrospective analysis from a tertiary academic memory disorders center, which may enrich for diagnostically complex referrals and limit generalizability to primary care or community neurology settings. Second, final diagnoses were biomarker-informed but were not uniformly validated against amyloid PET, CSF biomarkers, or neuropathology, limiting conclusions regarding absolute diagnostic correctness. Third, AD and non-AD classifications were simplified for reclassification analyses, whereas real-world neurodegenerative syndromes often involve mixed or overlapping pathologies. Finally, plasma ptau217 represents one component of an evolving biomarker ecosystem, and future comparative studies incorporating amyloid PET, CSF, tau PET, and longitudinal outcomes will be essential.

Despite these limitations, this study provides practical evidence that plasma p-tau217 meaningfully influences diagnostic decision-making in memory clinic practice, particularly among *APOE ε4* non-carriers and under threshold-dependent frameworks. Beyond diagnostic validity, the true value of blood-based biomarkers may lie in their ability to recalibrate clinical certainty, refine differential diagnosis, and better align patients with appropriate counseling, prognostication, and emerging disease-modifying therapies.^22–23^ As precision neurology advances, understanding not only whether biomarkers are accurate, but how they reshape real-world diagnostic behavior, will be critical.

## Disclosures

All authors reported no conflict of interest and financial disclosure.

| **Name** | **Location** | **Contribution** |
| --- | --- | --- |
| **Nora Maerean, MD** | Alzheimer's Disease and Memory Disorders Center, Department of Neurology, Baylor College of Medicine, Houston, TX | Drafting/revision of the manuscript for content, including medical writing for content; major role in collection of data, study design, analysis, and interpretation of data |
| **Sofia Litchev, BS** | Alzheimer's Disease and Memory Disorders Center, Department of Neurology, Baylor College of Medicine, Houston, TX | Drafting/revision of the manuscript for content, including medical writing for content; analysis and interpretation of data |
| **George R. Jackson, MD, PhD** | Alzheimer's Disease and Memory Disorders Center, Department of Neurology, Baylor College of Medicine, Houston, TX | Drafting/revision of the manuscript for content, including medical writing for content; analysis and interpretation of data |
| **Joseph S. Kass, MD, JD** | Alzheimer's Disease and Memory Disorders Center, Department of Neurology, Baylor College of Medicine, Houston, TX | Drafting/revision of the manuscript for content, including medical writing for content; analysis and interpretation of data |
| **Valory Pavlik, PhD** | Alzheimer's Disease and Memory Disorders Center, Department of Neurology, Baylor College of Medicine, Houston, TX | Drafting/revision of the manuscript for content, including medical writing for content; analysis and interpretation of data |
| **Chi-Ying R. Lin, MD, MPH** | Alzheimer's Disease and Memory Disorders Center, Department of Neurology, Baylor College of Medicine, Houston, TX | Drafting/revision of the manuscript for content, including medical writing for content; major role in collection of data, study design, analysis, and interpretation of data |

## References

1. Weintraub S, Wicklund AH, Salmon DP. The neuropsychological profile of Alzheimer Disease. Cold Spring Harb Perspect Med. 2012;2(4):a006171. doi: 10.1101/cshperspect.a006171.

2. Jack Jr CR, Andrews JS, Beach TG, et al. Revised criteria for diagnosis and staging of Alzheimer’s disease: Alzheimer’s Association Workgroup. Alzheimer’s & Dementia. 2024;20(8):5143–5169. doi: 10.1002/alz.13859.

3. Wei H, Masurkar AV, Razavian N. On gaps of clinical diagnosis of dementia subtypes: A study of Alzheimer’s disease and Lewy body disease. Front Aging Neurosci. 2023;15:1149036. doi: 10.3389/fnahi.2023.1149036.

4. Giebel C, Silva-Ribero W, Watson J, et al. A systemic review on the evidence of misdiagnosis in dementia on accessing dementia care. Int J Geriatr Psyciatry. 2024;39(10):e158. doi: 10.1002/gps.6158.

5. Rapaka D, Saniotis A, Mohammadi K, et al. From ambiguity to accuracy: A review of Alzheimer’s disease diagnostic errors and the need for non-invasive biomarkers. J Neurosci Rural Pract. 2025;16(1):14–21. doi: 10.25259/JNRP_431_2024

6. Wiedermann CJ, Piccoliori G, Engl A, von Strobele-Prainsack DH. Predictive biomarkers for asymptomatic adults: Opportunities, risks, and guidance for general practice. Diagnostics. 2026;16(2):196. doi: 10.3390/diagnostics16020196.

7. Augustinack JC, Schneider A, Mandelkow EM, Hyman BT. Specific tau phosphorylation sites correlate with severity of neuronal cytopathology in Alzheimer’s disease. Acta Neuropathol. 2002;103(1):26–35. doi: 10.1007/s004010100423.

8. Arranz J, Zhu N, Rubio-Guerra S, et al. Diagnostic performance of plasma pTau_217_, pTau_181_, Aβ_1-42_ and Aβ_1-40_ in the LUMIPULSE automated platform for the detection of Alzheimer disease. Alzheimer’s Res Ther. 2024;16(1):139. doi: 10.1186/s13195-024-01513-9.

9. Cecchetti G, Agosta F, Rugarli G, et al. Diagnostic accuracy of automated Lumipulse plasma pTau-217 in Alzheimer’s disease: a real-world study. J Neurol. 2024;271(10):6739–6749. doi: 10.1007/s00415-024-12631-7

10. Kumar M, Schlaffner CN, Tang S, et al. Molecular features of human pathological tau distinguish tauopathy-associated dementias. Cell. 2026;189(3):956–968.e13. doi: 10.1016/j.cell.2025.12.036.

11. Meyer MR, Kirmess KM, Eastwood S, et al. Clinical validation of the Precivity AD2 blood test: A mass spectrometry-based test with algorithm combining %p-tau217 and Aβ42/40 ratio to identify presence of brain amyloid. Alzheimers Dement. 2024;20(5):3179–3192. doi: 10.1002/alz.13764.

12. Ashton NJ, Brum WS, Di Molfetta G, et al. Diagnostic accuracy of a plasma phosphorylated tau 217 immunoassay for Alzheimer’s disease pathology. JAMA Neurol. 2024;81(3):255–263. doi: 10.1001/jamaneurol.2023.5319.

13. McKhann G, Drachman D, Folstein M, Katzman R, Price D, Stadlan EM. Clinical diagnosis of Alzheimer’s disease: report of the NINCDS-ADRDA Work Group under the auspices of Department of Health and Human Services Task Force on Alzheimer’s Disease. Neurology. 1984;34(7):939–944. doi:10.1212/wnl.34.7.939.

14. Labcorp. Phosphorylated Tau 217 (pTau-217), Plasma. Accessed June 4, 2026. https://www.labcorp.com/tests/484390/phosphorylated-tau-217-ptau-217-plasma

15. Figdore DJ, Griswold M, Bornhorst JA, et al. Optimizing cutpoints for clinical interpretation of brain amyloid status using plasma p-tau217 immunoassays. Alzheimers Dement. 2024;20(9):6506–6516. doi: 10.1002/alz.14140.

16. De Strooper B, Zetterberg H. Tracking the turning point in Alzheimer’s disease. Science. 2026;392(6797):468–469. doi: 10.1126/science.aeb6987.

17. Middleton LE, Grinberg LT, Miller B, Kawas C, Yaffe K. Neuropathologic features associated with Alzheimer disease diagnosis: age matters. Neurology. 2011;77(19):1737–1744. doi: 10.1212/WNL.0b013e318236f0cf.

18. Robinson JL, Richardson H, Xie SX, et. al. The development and convergence of co-pathologies in Alzheimer’s disease. Brain. 202l;144(3):953–962. doi: 10.1093/brain/awaa438.

19. Schneider JA, Arvanitakis Z, Bang W, Bennett DA. Mixed brain pathologies account for most dementia cases in community-dwelling older persons. Neurology. 2007;69(24):2197–2204. doi: 10.1212/01.wnl.0000271090.28148.24.

20. Hampel H, Hu Y, Cummings J, et al. Blood-based biomarkers for Alzheimer’s disease: Current state and future use in transformed global healthcare landscape. Neuron. 2023;111(18):2781–2799. doi: 10.1016/j.neuron.2023.05.017.

21. Slikker W. Biomarkers and their impact on precision medicine. Exp Biol Med (Maywood). 2017;243(3):211–212. doi: 10.1177/1535370217733426.

22. Cummings J, Apostolova L, Rabinovici GD, et al. Lecanemab: Appropriate use recommendations. J Prev Alzheimers Dis. 2023;10(3):362–377. doi: 10.14283/jpad.2023.30.

23. Rabinovici GD, Selkoe DJ, Schindler SE, et al. Donenemab: Appropriate use recommendations. J Prev Alzheimers Dis. 2025;12(5):100150. doi: 10.1016/j.tjpad.2025.100150.

