## Supplemental Table 1 for "Plasma Phosphorylated-tau 217 Reshapes Diagnostic Classification in a Real-World Memory Clinic Cohort"

**Supplemental Table 1.** Initial impressions and diagnoses that were unmatched among study participants

| **Participant** | **Initial impression** | **Current diagnosis** |
| --- | --- | --- |
| No. 1 | profound dementia due to VaD | profound dementia due to mixed AD and VaD |
| No. 2 | moderate dementia suspected due to DLB | moderate dementia due to AD and VaD |
| No. 3 | multidomain MCI due to psychiatric disorder, would benefit from AD testing | multidomain MCI due to AD pathology and mood disorders |
| No. 4 | forgetfulness likely resulting from ADHD symptoms | AD pathology based on ptau biomarker |
| No. 5 | amnestic multidomain MCI, less likely due to AD | amnestic mild MCI due to AD |
| No. 6 | nonfluent PPA due to FTD with mild dementia | mild dementia due to AD, parieto-occipital type |
| No. 7 | suspicion for early non-amnestic MCI, consideration for DLB | nonamnestic MCI due to AD |
| No. 8 | rapidly progressive dementia in setting of chronic alcoholism and anti-P/Q type voltage dependant Ca channel antibodies | mild to moderate dementia due to AD and VaD |
| No. 9 | suspicion for mild dementia due to bvFTD or frontal variant AD | moderate dementia due to vascular etiology |
| No. 10 | amnestic multi-domain MCI, suspected due to AD | mild dementia, etiology undeteremined FTD or AD |
| No. 11 | amnestic MCI due to AD | non-amnestic MCI captured by NPS, unlikely due to AD |
| No. 12 | amnestic MCI due to AD | does not meet criteria for neurocognitive disorders |
| No. 13 | amnestic MCI due to AD | non-amnestic MCI with etiology to be determined but per biomarker is not AD |
| No. 14 | worsening of original baseline cognitive impairment or dysexecutive/frontal variant AD | major neurocognitive disorder, suspected etiology NF-1 |
| No. 15 | mild to moderate dementia due to AD | moderate dementia with subcortical features identified via NPS and the brain MRI showed frontal and parietal predominant atrophy, instead of hippocampal atrophy. Precivity AD2 testing was negative. |
| No. 16 | amnestic MCI due to AD | mild severity of vascular dementia |
| No. 17 | mild-moderate dementia due to mixed AD and VaD | mild to moderate dementia due to VaD |
| No. 17 | MCI, most likely due to AD pathology | not meeting neurocognitive dx per NPS testing, ptau negative for AD pathology |
| No. 18 | amnestic MCI due to AD | amnestic MCI but no biomarker evidence for AD. Likely PART |
| No. 19 | mild dementia due to mixed AD and VaD | mild major neurocognitive disorder in NPS primarily driven by psychiatric disorder |
| No. 20 | early MCI, but multiple confounders including low Vit D and AD | doesn't meet criteria for neurocog disorder per NPS, AD biomarker testing negative |
| No. 21 | early amnestic MCI due to AD | doesn't meet criteria for neurocognitive disorder per NPS, low AD likelihood |
| No. 22 | amnestic MCI due to combined MS and AD | amnestic MCI, more likely due to MS given negative AD2 testing |
| No. 23 | amnestic MCI due to AD | non-amnestic MCI due to VaD |
| No. 24 | early amnestic MCI due to AD | not meeting criteria for neurocognitive disorder per neuropsychological testing; etiology-wise, his brain MRI raises concern for AD change based on the cerebral volume loss and cortical microhemorrhage, but Precivity AD2 returned negative for AD |
| No. 25 | moderate dementia due to AD | moderate dementia less likely due to AD with negative ptau217, second most likely diagnosis is FTD |
| No. 26 | amnestic multidomain MCI due to AD | subthreshold MCI due to vascular etiology; likelihood of AD is now low due to precivity AD2 results |
| No. 27 | early amnestic MCI due to AD | unlikely AD due to low Precivity AD2 |
| No. 28 | multidomain MCI due to post cortical pathology indicative of AD pathology | frontotemporal degeneration; negative AD2 biomarker, frontal lobe dysfunction on FDG-PET |
| No. 29 | amnestic mild cognitive impairment due to AD | amnestic MCI etiology to be determined, consider FTD |
